# Tracking the spread of novel coronavirus (2019-nCoV) based on big data

**DOI:** 10.1101/2020.02.07.20021196

**Authors:** Xumao Zhao, Xiang Liu, Xinhai Li

## Abstract

The novel coronavirus (2019-nCoV) appeared in Wuhan in late 2019 have infected 34,598 people, and killed 723 among them until 8^th^ February 2020. The new virus has spread to at least 316 cities (until 1^st^ February 2020) in China. We used the traffic flow data from Baidu Map, and number of air passengers who left Wuhan from 1^st^ January to 26^th^ January, to quantify the potential infectious people. We developed multiple linear models with local population and air passengers as predicted variables to explain the variance of confirmed cases in every city across China. We found the contribution of air passengers from Wuhan was decreasing gradually, but the effect of local population was increasing, indicating the trend of local transmission. However, the increase of local transmission is slow during the early stage of novel coronavirus, due to the super strict control measures carried out by government agents and communities.

## Introduction

Since the first case of pneumonia named 2019 Novel Coronavirus (2019-nCoV) being identified in in Wuhan, China, a total of 34,598 confirmed infections were reported in the country until 8^th^ February 2020, which caused 723 deaths (1). Guo et al (2020) have found that 2019-nCoVs was similar with bat coronaviruses (2), which occasionally transmitted to human due to wildlife consumption. Understanding the pattern of transmission characteristics of 2019-nCoV is important for effective preventing and controlling the ongoing pandemic disease. The value of *R*_*0*_ (basic reproduction number) was estimated as 2.2, inferring a median size outbreak (3). However, based on the epidemic transmission model, the number of actual infections would be much larger than the number of confirmed cases (4).

At present, tracking the passengers from Wuhan in January 2020 is still the top task for preventing the further spread of novel coronavirus (2019-nCoV). To accurately estimate the risk of the novel coronavirus, we compiled the detailed daily traffic data outbound Wuhan from a big-data source, Baidu Map, before the lockdown of Hubei Province, in order to provide information for risk assessment of 2019-nCoV at the province level and the city level (Supp. Fig.1).

## Methods

The traffic flow data outbound Wuhan from 1^st^ January to 26^th^ January 2020 was downed from Baidu Map Huiyan platform (5). The number of air passengers from Wuhan from 30^th^ December 2019 to 20 January 2020 was released by Aviationtalk (6). The time series data of confirmed 2019-nCoV cases from 10^th^ January to 30 January 2020 was obtained from People’s daily-Dingxiangyuan (1), which was released by China National Health Commission.

We did Spearman correlation analysis for the daily traffic from Wuhan (from 1^st^ January to 26^th^ January) and the total traffic in this period with the number of confirmed cases (from 25^th^ January to 30^th^ January). To explain the variance of confirmed cases in all infected provinces, we developed multiple linear models including population, GDP, population density, and mean temperature as independent variables. All analysis was performed using R (version 3.6.2).

## Results

From 20^th^ December 2019 to 20 January 2020, 854,424 air passengers left Wuhan Tianhe Airport to 49 cities in China (Fig. 1). From 1^st^ to 26^th^ January, about three million domestic passengers travelled from Wuhan to other cities. Among the passengers, a few thousands had been confirmed to infected by the novel coronavirus (Fig. 2). The distribution of air passengers from Wuhan to other cities in China had high correlation coefficients (0.71) with the number of confirmed infection cases in those cities on 22^nd^ January. The correlation coefficient drops to 0.56 on 24^th^ January. Then the number of confirmed infection cases was positive correlation with local population size.

**Fig. 1.**
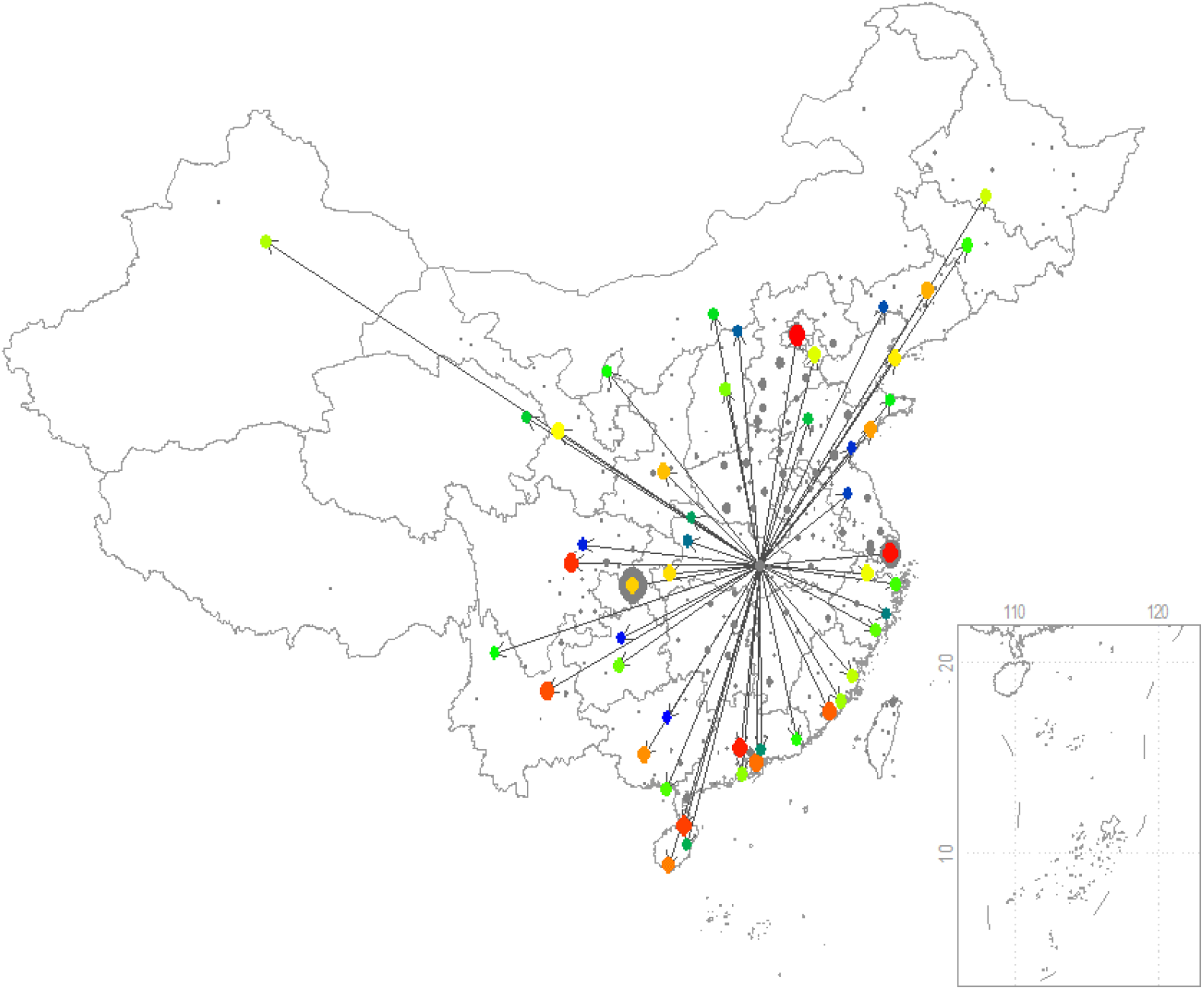
The distribution of 854,424 air passengers from Wuhan Tianhe Airport to 49 cities in China during the period from 30^th^ December 2019 to 20^th^ January 2020. The sizes of the points indicate the number of passengers, which are also represented in color series: blue, green, yellow, brown, and red.

**Fig. 2.**
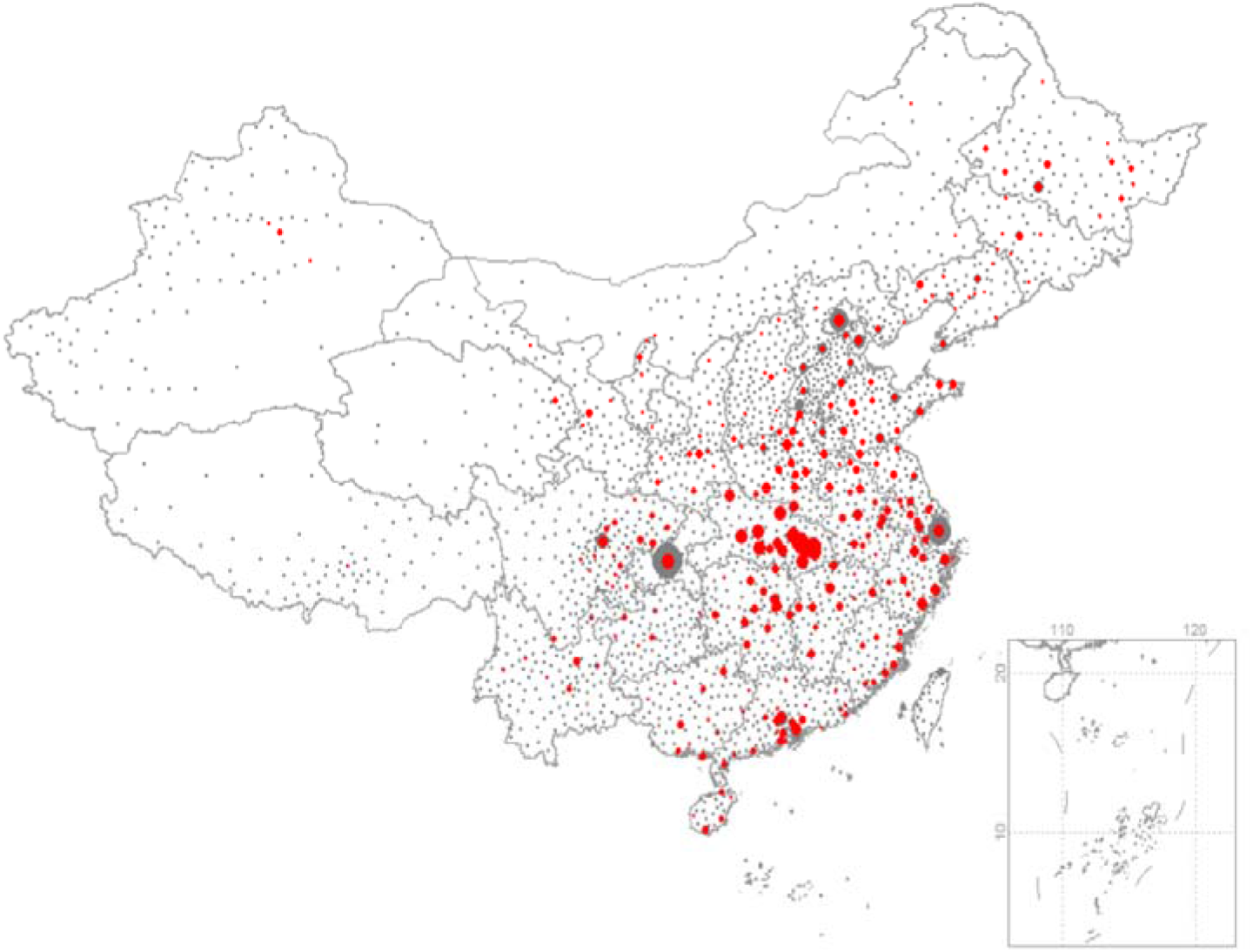
The spatial distribution of 24,281 confirmed cases (incomplete data) of 2019-nCoV on 5^th^ February in 299 cities in China. The sizes of red points represent the number of confirmed cases. The sizes of grey points show the population of the cities.

We used a multiple regression model to explain the variance of the number of cases in the infected cities. After model selection, only two variables remained, the number of passengers and local population (Fig. 3). Overall, the population of the provinces explains near half of the variance in the number of confirmed cases across 34 provinces and province-level municipalities, whereas the number of passengers from Wuhan explained around 10% (Fig. 3).

**Fig. 3.**
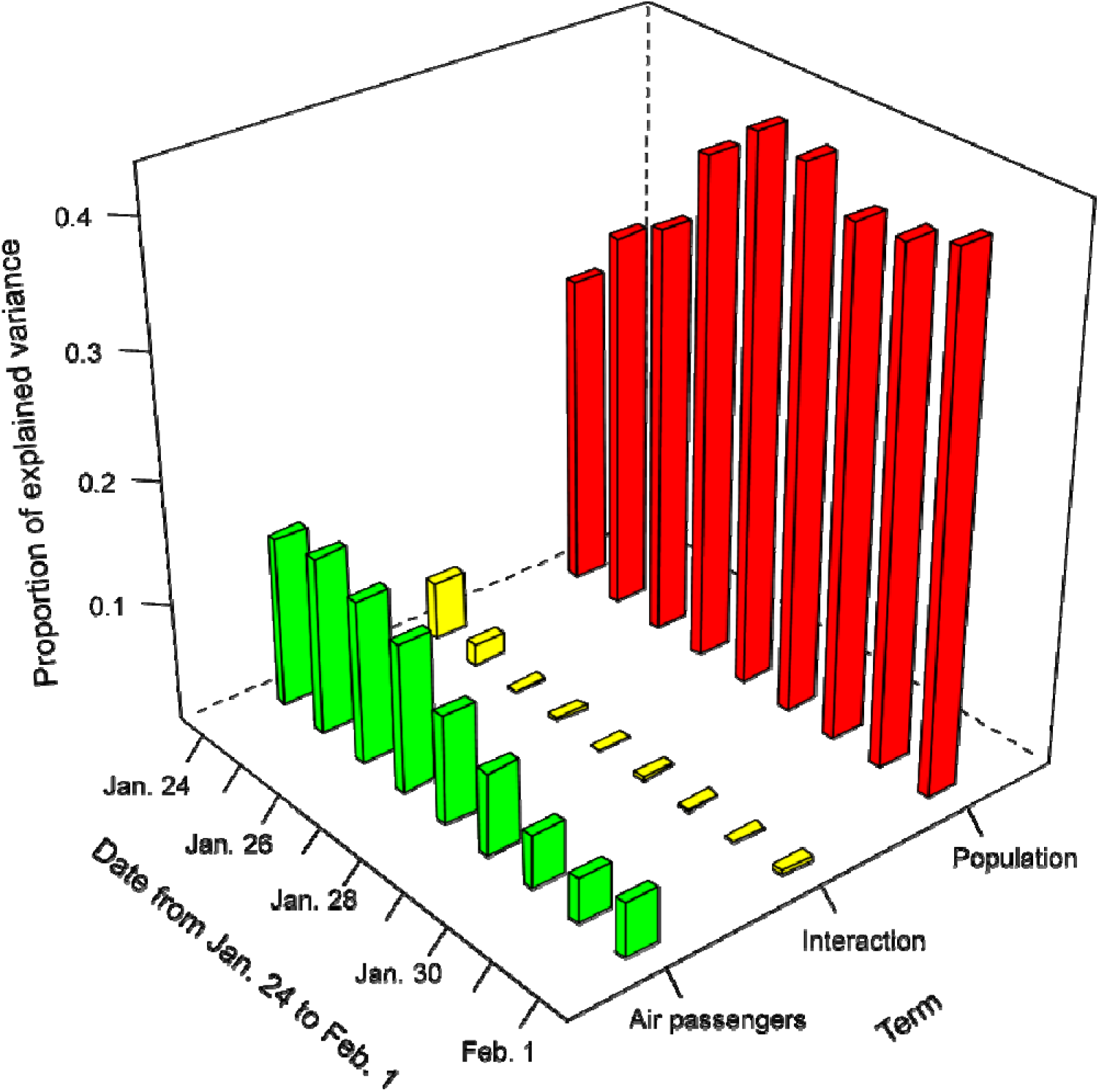
The proportion of explained variance in linear regression: Number of confirmed cases in 34 provinces ∼ Air passengers from Wuhan * Population of the province for the date from 24^th^ January to 1^st^ February.

Correlation coefficients of number of confirmed that the number of cases in the cities (n=97) from 25^th^ January to 30^th^ January match the number of passengers from Wuhan during the period from 1^st^ January to 26^th^ January. The highest correlation appears on 5^th^ January, inferring a long incubation period up to two weeks (Supp. Table 1).

## Discussion

In the beginning of the spread of the pneumonia, there is a high correlation (0.71) between the number of confirmed infection cases and air passengers from Wuhan, proofed Wuhan the source of the pneumonia (7). As time going, local population played a more dominant role, because the local spread of 2019-nCoV is likely to happen. The basic reproductive number of the infection (*R*_0_) to be estimated as 3.8 means 72-75% of transmissions must be prevented to stop the outbreak (4). Fortunately, the transmission of the virus was really controlled due to strict prevention measures carried out by Chinese government. Restricted population movements ban was enforced upon 16 cities in Hubei Province since 23^rd^ January 2020 (8), resulting in significant decrease in passengers from Wuhan and adjacent cities, which effectively reduces the spread of the pneumonia. However, 3-5 million people had left Wuhan for numerous cities in China before the province lockdown (Supp. Fig.2), and among them a number of infected people have no clinical symptom yet infectious to others. We believe this is the highest challenge against the current national level antivirus campaign.

Currently the first-level response to major public health emergencies has been initiated in 30 provinces, municipalities and autonomous regions in China on 25^th^ January 2020 (9), so that strict control procedures are carried out to prevent the spread of the virus. According to an infectious disease model, it was estimated that the actual number of infected people in Wuhan on 4^th^ February 2020 may reach 250 thousand (prediction interval, 164,602 to 351,396) without any control (3). Whereas only 5,142 confirmed infections were reported in China on 2^nd^ February 2020. Starting from 3^rd^ February, all suspected case in Wuhan will be taken in to medical care, and the virus spread in Wuhan can be controlled gradually.

The correlation coefficients between the number of confirmed cases from 26^th^ January to 30^th^ January and number of passengers from Wuhan were higher than that on 25^th^ January (Supp. Table 1). We think the reason is that the confirmed cases on 25^th^ is too low due to lack of virus detection kit. After 25^th^ January the supply of virus detection kit was enough and the number of confirmed cases reflect the real situation, which have very high correlation with the number of passengers from Wuhan to these cities during 1^st^ January to 26^th^ January. We notice the highest correlation appear on 5^th^ January, two to three weeks ahead of the confirmed cases in those cities (confirmation also needs several days to complete at that time), which infers the long latent period of 2019-nCoV.

Combining the two variables (local population and passengers from Wuhan) to interpret the virus outbreak risk, we recommend that the preventing and controlling measures can be divided into two different stages. In the early stage, checking the passengers from Wuhan is more important. Some cities (e.g., Wenzhou in Zhejiang Province) with a large number of returnees from Wuhan have many cases even when they are far from Wuhan in space. The database of travel routes of confirmed patients has been developed and published for free use (10), in order to tracking and warning close contactors. In the late stage, local population become the most important factor in predicting the number of confirmed infection cases. This suggests us that the densely populated metropolitan areas, such as Shanghai, Beijing, Guangzhou, and Shenzhen should pay special attentions to preventing the second-generation infections. Moreover, the densely populated rural areas around Wuhan may face double threats of the spread of infectious people from Wuhan and a large number of local susceptible people. These areas, including areas of Hubei Province except for Wuhan, and surrounding area of Henan (Nanyang, Xinyang) and Chongqing, need to prepare for a surge in infection. The rural medical facilities in these areas are scarcer than in cities, clustering cases are more likely to happen in these places.

## Data Availability

The data used in the manuscript were posted at the author's Githab account and freely available to public.

https://github.com/Xinhai-Li/2019-nCoV/blob/master/2019-NCoV_cities.xlsx

## Supplementary documents

**Supp. Fig. 1.**
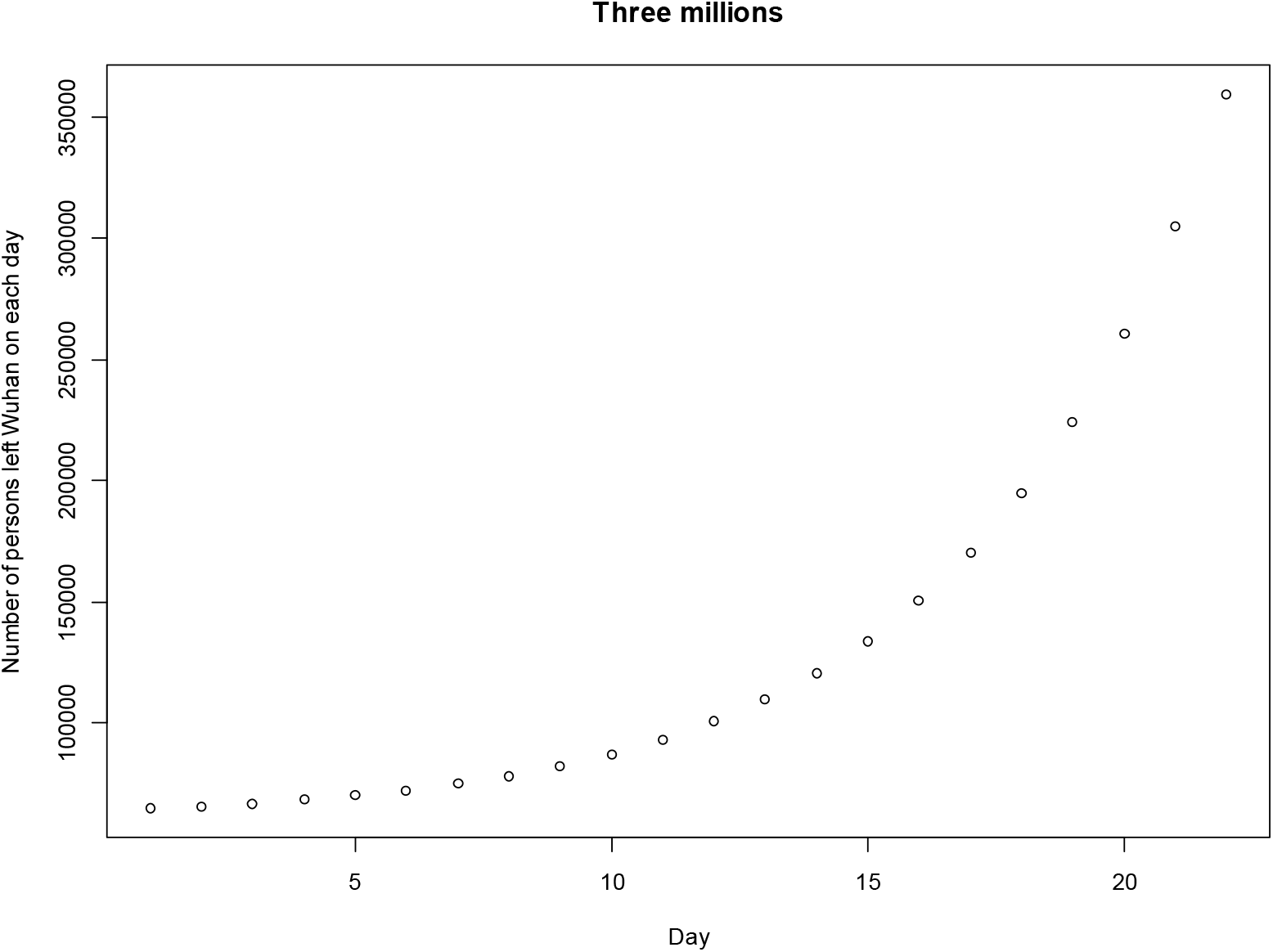
The daily passenger (include all vehicles) numbers from Wuhan to top 50 cities in China from 1st January to 26th January are showed by an animated GIF file 2019-nCoV_spread_1-26 Jan.gif (Supp. Fig. 1).

**Supp. Fig. 2.**
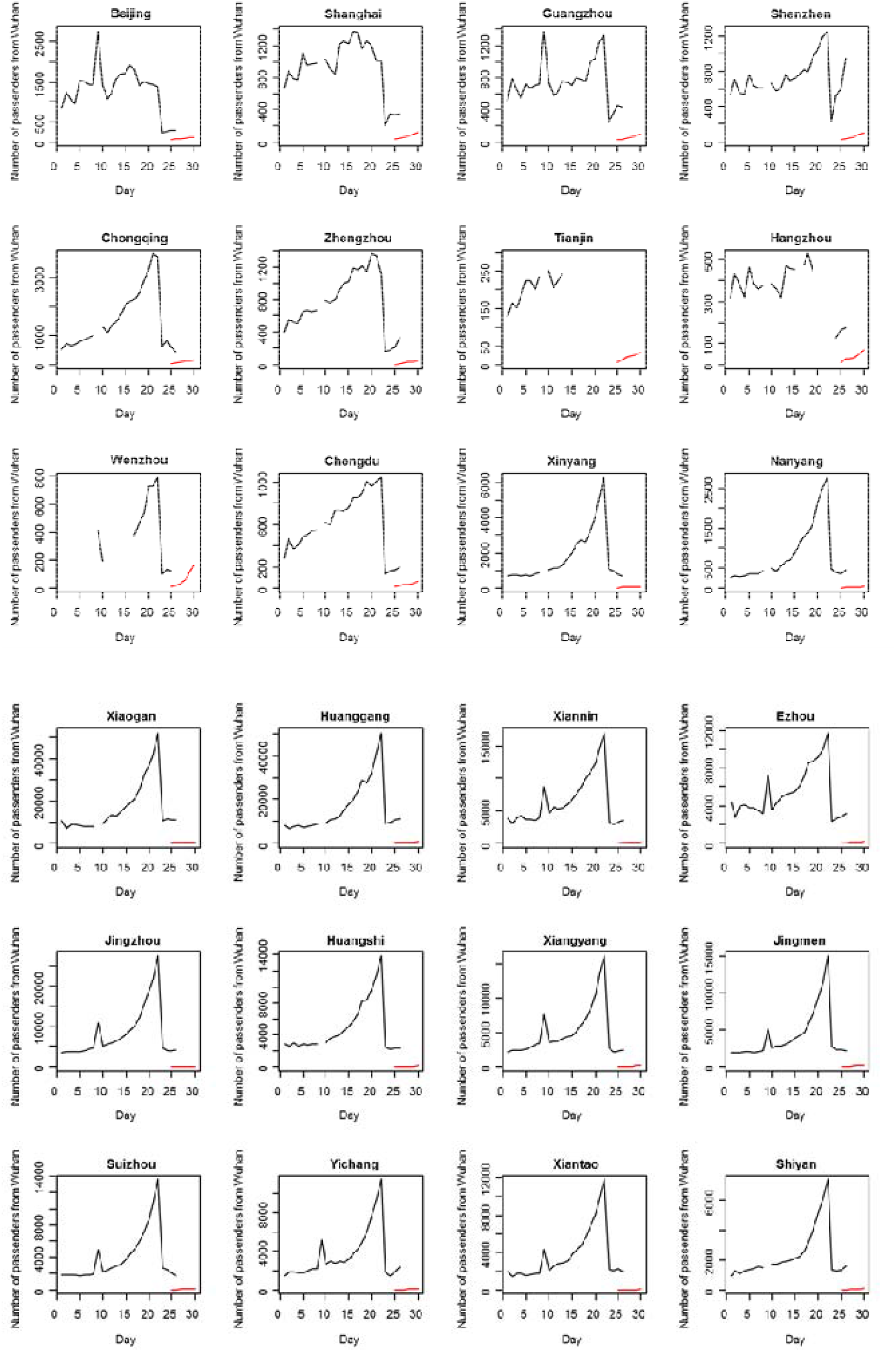
Estimated daily number of passengers from Wuhan using a logistic curve y = 60000 + 10000/exp(1-0.2*x) from 1^st^ January to 22^nd^ January (the day before city lockdown). The assumption is that people tend to leave Wuhan just before the Chinese New Year on 25^th^ January.

**Supp. Fig. 3.**
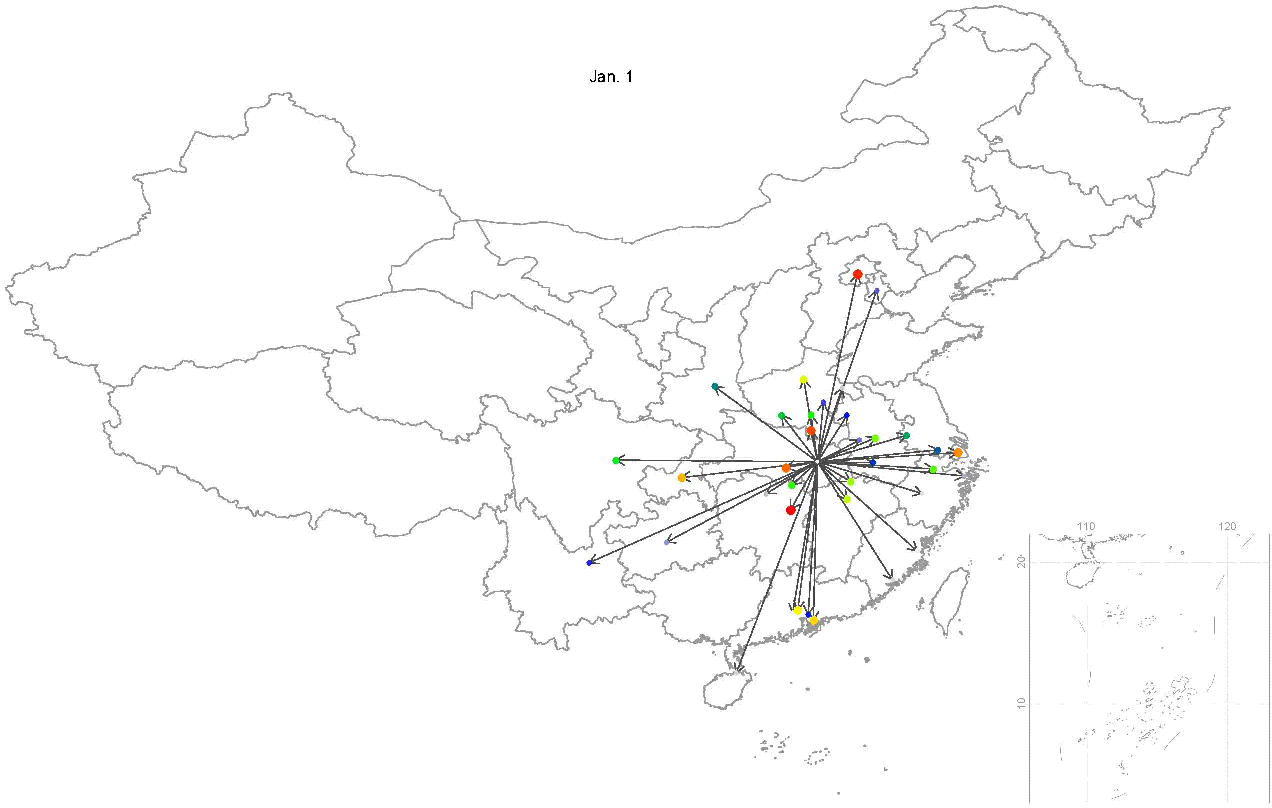
Estimated daily number of passengers from Wuhan to 12 major cities in China (upper panel) and 12 major cities in Hubei Province (lower panel) base on Baidu Bigdata server. The red lines show the daily accumulated confirmed case in each city from 25^th^ January to 30^th^ January.

